# House value as an individual socioeconomic indicator for breast cancer survival and late-stage diagnosis: a population-based cohort study from Northern Ireland

**DOI:** 10.1101/2025.10.13.25337879

**Authors:** Sarah Baxter, Charlene M. McShane, Stuart A. McIntosh, Damien Bennett, Meenakshi Sharma, Lynne Lohfeld, Daniel R.S. Middleton, Gerard Savage, Deirdre Fitzpatrick, Ann Mc Brien, David McCallion, Anna Gavin, Chris R. Cardwell

## Abstract

**Background:** Socio-economic inequalities in breast cancer survival persist globally, including in the UK. Area-based deprivation measures may underestimate true socio-economic effects by assigning average levels to all individuals within an area. This study investigated associations between house value (individual-level socio-economic indicator) and area-based deprivation with breast cancer outcomes in Northern Ireland.

**Methods:** Women diagnosed with breast cancer 2011 to 2021 were identified using the Northern Ireland Cancer Registry. House value was determined from Valuation and Lands Agency property valuation data, and area-based deprivation was determined from the Northern Ireland Multiple Deprivation Measure. The primary outcome was breast cancer-specific mortality. Secondary outcomes included stage at diagnosis. Cox regression models calculated adjusted hazard ratios (HR) and (95%CIs) for cancer-specific mortality by house value category and separately for deprivation, adjusting for confounders.

**Results:** Among 12,766 women with breast cancer, associations were much more pronounced for house value than area-based deprivation. Women in the lowest house value category, compared to the highest value category, had a 60% increase in mortality (adjusted HR=1.60 95%CI 1.34, 1.92) and were more likely to be diagnosed with stage 4 disease (7.5% versus 4.1%; P<0.001). Women living in the most versus least deprived areas had a 26% increase in mortality (adjusted HR=1.26 95%CI 1.08, 1.47) and were more likely to be diagnosed with stage 4 disease (5.9% vs 5.0%; P =0.157).

**Conclusion:** House value demonstrated stronger associations with breast cancer outcomes than area-level deprivation, suggesting it may serve as a more sensitive indicator for monitoring health inequalities in cancer.

**What is already known:** Lower socioeconomic status is associated with worse breast cancer survival in the UK and internationally. Area-based measures of deprivation are widely used in epidemiological research but have recognised limitations.

**What this study adds:** We observed socio-economic gradients for both late-stage at diagnosis and breast cancer survival for both house value and area-deprivation. However, associations were stronger for house value and effects persisted after adjustment for area deprivation.

**How this might affect research, policy and practice:** Our findings establish that publicly available property valuations provide a feasible and sensitive measure for monitoring socioeconomic health inequalities at the population level

## Introduction

Breast cancer is the most commonly diagnosed cancer among women worldwide, with over 2.3 million cases in 2022 [1]. Although breast cancer survival has improved in recent decades [2, 3], breast cancer patients from lower socio-economic backgrounds have demonstrated poorer survival, both in the UK [4, 5] and internationally [6–8]. Despite efforts to reduce socio-economic inequalities in cancer outcomes within the UK, these disparities persist [9–11].

The majority of research on socio-economic inequalities in breast cancer mortality has focused on area-based measures of deprivation [5], primarily because cancer registries do not capture information on individual-level measures of socio-economic status (SES). Area-level measures of deprivation are calculated by allocating individuals to administrative geographical areas and aggregating indicators across these areas. This approach offers advantages such as deprivation data are typically publicly available for the entire population, and these measures can capture different domains of deprivation.

However, area-level measures have recognised limitations. Most importantly, they assign individuals the average deprivation level of their area, which reduces the scale of inequalities towards the null [12]. Additionally, area-based and individual-level measures often classify patients, including cancer patients [13], differently. For instance, affluent individuals living in deprived areas are classified according to their area-level rather than individual SES, and vice versa. This exposure misclassification increases as administrative geographies become larger [12].

Investigating individual-level SES is important to better understand the mechanisms underlying health inequalities [13]. House value, the estimated monetary worth of a residential property, has been proposed as a novel individual-level SES measure that reflect both economic resources and neighbourhood context [14, 15]. House value may be particularly valuable for older populations who may have retired, making traditional occupation-based or income-based SES measures less relevant [14]. House value may be especially relevant in the UK context, where homeownership rates are high [16] and property wealth constitutes a substantial portion of household assets [14, 17, 18].

Despite this, few studies have previously investigated house value and mortality in breast cancer patients [19, 20]. In Northern Ireland (NI) the house valuation for all domestic properties is publicly available [14], making this a potentially useful population-level SES measure. Consequently, we examined the association between individual-level house value and area-based deprivation with breast cancer-specific mortality and stage at diagnosis in a large cohort of breast cancer patients from NI.

## Methods

### Data sources

The study utilised data from the Northern Ireland Cancer Registry (NICR), which captures data on all patients diagnosed with cancer in NI. The NICR contains comprehensive information obtained from the Health and Social Care Northern Ireland (HSCNI) including age, date of diagnosis, tumour site, tumour stage and grade, as well as death registrations from the General Registrar Office. The NICR maintains high accuracy with an estimated completeness exceeding 99% [21, 22]. Patient residential addresses at cancer diagnosis were obtained from the Northern Ireland Health Card registration system and linked to property valuations from the Valuation and Lands Agency Northern Ireland-wide government property valuation exercise using unique property identifiers. Details of this linkage process are provided in the exposure section below.

Prescription medication data were taken from the Northern Ireland Electronic Prescribing Dataset (NIEPD), maintained by the Health and Social Care Business Services Organisation, which captures all NHS prescriptions dispensed by community pharmacists in NI. All datasets were linked using linked using a unique patient identifier (NI Health and Care Number) by HSCNI Honest Broker Service [23] . All analyses were conducted within the Honest Broker Service secure environment. Ethical approval for this study was obtained from Bradford Leeds Research Ethics Committee (reference: 23/YH/0157).

### Cohort

From the NICR we identified all women newly diagnosed with invasive breast cancer (ICD-10 code C50) between January 2011 and December 2021. Women with a previous diagnosis of any cancer (apart from non-melanoma skin cancer) were excluded.

### Outcome

The primary outcome was breast cancer-specific mortality, defined as death with breast cancer (ICD-10 code C50) as the underlying cause of death. Secondary outcomes were stage 4 at diagnosis and all-cause mortality. In sensitivity analyses, we examined both stage 4 and stage unknown at diagnosis.

### Exposure

The exposure variables of interest were house value of patients’ residence at time of breast cancer diagnosis (individual-level SES measure) and the 2017 Northern Ireland Multiple Deprivation Measure [24] (NIMDM) classification (area-level classification).

To determine house value, patient addresses at diagnosis were obtained from the Northern Ireland Health Card registration system, which maintains addresses for all patients accessing NHS healthcare and has been estimated to have high completeness [25]. Each address was matched to its Unique Property Reference Number (UPRN) using the Pointer address database maintained by Land and Property Services. The ratable house value, which is still used in 2025 to determine local tax levels, was obtained and was based upon a 2007 valuation exercise undertaken by the former Valuation and Lands Agency in which all domestic properties were valued according to their estimated open market value as of 2005. Properties built after 2005 were assigned values equivalent to comparable pre-2005 properties [26]. House values were available in eight categories (in £1,000s): <75, 75-99, 100-124, 125-199, 200-249, 250-299, 300-399, and ≥400. For this analysis, house values were classified into the following five categories (in £1000s): <75, 75-99, 100-124, 125-199, ≥200.

The NIMDM was determined for each patient’s postcode at diagnosis, based upon Super Output Areas (geographic units of approximately 2,100 people). The NIMDM captures seven domains of deprivation: income, employment, health, education, access to services, living environment and crime, and was analysed in fifths.

### Covariates

The NICR provided information on year of diagnosis, age at diagnosis, tumour stage and grade, and pathway to diagnosis (screen detected, red flag referral, death certificate only, and other). Treatment data within the first year of diagnosis included surgery (mastectomy or breast-conservation), systemic therapy (including chemotherapy) and radiotherapy. Use of endocrine treatments (tamoxifen or aromatase inhibitors) was determined from dispensed medications recorded in the NIEPD.

Pre-existing comorbidities were identified from hospital admissions data up to five years prior to breast cancer diagnosis. The following conditions from the Charlson Comorbidity Index were identified based on ICD-10 codes as a cause of hospital admission from Patient Administration System, using previously used code lists [27]. The conditions included: myocardial infarction, congestive heart disease, peripheral vascular disease, cerebrovascular disease, chronic pulmonary disease, dementia, liver disease, peptic ulcer disease, diabetes and chronic kidney disease.

### Statistical analysis

First, characteristics of patients with breast cancer were determined by house value. Cox regression analysis was used to examine associations between SES measures and breast cancer-specific mortality. Follow-up was censored at death from other causes, date of emigration and date of complete mortality records (31^st^ March 2023). Kaplan-Meier curves were plotted to show survival by house value. Three levels of adjustment were applied: (1) age at diagnosis, year of diagnosis, individual Charlson comorbidities; (2) additionally including area deprivation; and (3) additionally including tumour stage and grade. Analyses were repeated with area deprivation as the exposure.

The main analysis used a complete case approach where we excluded patients with missing house value or deprivation. However, we assessed the impact of missing data by conducting two further sensitivity analyses. First, we included a missing category for house value and deprivation. Second, we conducted an analysis using multiple imputation for missing house value and deprivation. House value and deprivation (in fifths) were imputed, using chained equations, in 10 datasets using ordinal logistic regression models with cancer-specific death status, cumulative hazard and age at diagnosis, year of diagnosis and Charlson comorbidities in imputation models [28], and results were combined using Rubin’s rules [29].

Logistic regression analysis was used to examine associations between SES measures and stage 4 disease at diagnosis. In the primary analysis we compared stage 4 disease to stage 1-3 disease at diagnosis. Models were fitted separately for house value and area deprivation as exposures. Adjusted analysis included age at diagnosis, year of diagnosis, and individual Charlson comorbidities, with area deprivation additionally included in house value models. In sensitivity analysis we examined stage 4 or stage unknown disease at diagnosis versus stage 1 to 3 disease at diagnosis. Again, we utilised a complete case approach (those with either a missing house value or deprivation were excluded). All analyses were conducted using STATA 18 (StataCorp, Texas, USA).

## Results

The full cohort included 13,846 women with breast cancer. Overall, 976 (7%) women had a missing house value and 104 (1%) had a missing deprivation score, leaving 12,766 women in the main analysis with complete records for house value and deprivation.

### Characteristics of patients

Characteristics of patients by house value are shown in Table 1, demonstrating clear socio-economic gradients across several characteristics. Compared with patients living in the most valuable properties, those living in the least valuable properties were slightly older, had slightly higher stage, were more likely to be detected via red flag referral, and were less likely to have radiotherapy, systemic therapy and be prescribed tamoxifen. There were clear differences in area-based deprivation by house value, with only 39% of breast cancer patients in the least valuable properties living in the most deprived areas, and only 45% of patients in the most valuable properties living in the least deprived areas. Other characteristics were similar across house value categories.

**Table 1.** Characteristics of breast cancer patients by house value.

| Characteristics | House value (per £1000) |  |  |  |  |
| --- | --- | --- | --- | --- | --- |
|  | 0-74 | 75-99 | 100-124 | 125-199 | >200 |
| *n* | 2,483 | 2,608 | 2,118 | 3,722 | 1,835 |
| Year: 2010-13 | 669 (27%) | 695 (27%) | 528 (25%) | 901 (24%) | 432 (24%) |
| 2014-16 | 647 (26%) | 658 (25%) | 583 (28%) | 1058 (28%) | 495 (27%) |
| 2017-19 | 674 (27%) | 759 (29%) | 608 (29%) | 1076 (29%) | 556 (30%) |
| 2020-22 | 493 (20%) | 496 (19%) | 399 (19%) | 687 (18%) | 352 (19%) |
| Age: <50 | 436 (18%) | 536 (21%) | 478 (23%) | 766 (21%) | 475 (26%) |
| 50-<60 | 530 (21%) | 646 (25%) | 544 (26%) | 985 (26%) | 525 (29%) |
| 60-<70 | 638 (26%) | 635 (24%) | 516 (24%) | 945 (25%) | 440 (24%) |
| 70-<80 | 492 (20%) | 467 (18%) | 358 (17%) | 623 (17%) | 250 (14%) |
| 80+ | 387 (16%) | 324 (12%) | 222 (10%) | 403 (11%) | 145 (8%) |
| Deprivation (in fifths): |  |  |  |  |  |
| 1<sup>st</sup> (Most deprived) | 964 (39%) | 673 (26%) | 250 (12%) | 219 (6%) | 45 (2%) |
| 2<sup>nd</sup> fifth | 673 (27%) | 642 (25%) | 436 (21%) | 605 (16%) | 183 (10%) |
| 3<sup>rd</sup> fifth | 474 (19%) | 568 (22%) | 453 (21%) | 733 (20%) | 275 (15%) |
| 4<sup>th</sup> fifth | 283 (11%) | 452 (17%) | 535 (25%) | 1003 (27%) | 507 (28%) |
| 5<sup>th</sup> (least deprived) | 89 (4%) | 273 (10%) | 444 (21%) | 1162 (31%) | 825 (45%) |
| Stage: 1 | 936 (38%) | 1007 (39%) | 855 (40%) | 1562 (42%) | 758 (41%) |
| 2 | 945 (38%) | 1007 (39%) | 809 (38%) | 1452 (39%) | 723 (39%) |
| 3 | 306 (12%) | 330 (13%) | 288 (14%) | 425 (11%) | 224 (12%) |
| 4 | 177 (7%) | 146 (6%) | 96 (5%) | 163 (4%) | 73 (4%) |
| Missing | 119 (5%) | 118 (5%) | 70 (3%) | 120 (3%) | 57 (3%) |
| Grade: 1 | 296 (12%) | 280 (11%) | 233 (11%) | 435 (12%) | 227 (12%) |
| 2 | 1140 (46%) | 1220 (47%) | 1016 (48%) | 1777 (48%) | 897 (49%) |
| 3 | 898 (36%) | 989 (38%) | 795 (38%) | 1357 (36%) | 656 (36%) |
| Missing | 149 (6%) | 119 (5%) | 74 (3%) | 153 (4%) | 55 (3%) |
| Pathway: Screen detected | 669 (27%) | 766 (29%) | 625 (30%) | 1189 (32%) | 547 (30%) |
| Red flag referral | 1385 (56%) | 1390 (53%) | 1140 (54%) | 1803 (48%) | 826 (45%) |
| Death cert only | 0-10 (0%) | 0-10 (%) | 0-10 (%) | 0-10 (%) | 0-10 (%) |
| Other | 419-429<sup>a</sup> (17%) | 442-452<sup>a</sup> (%) | 343-353<sup>a</sup> (%) | 720-730<sup>a</sup> (%) | 452-462<sup>a</sup> (%) |
| Mastectomy | 669 (27%) | 732 (28%) | 551 (26%) | 960 (26%) | 421 (23%) |
| Breast-conservation | 1305 (53%) | 1392 (53%) | 1230 (58%) | 2060 (55%) | 1008 (55%) |
| Radiotherapy | 1654 (67%) | 1851 (71%) | 1567 (74%) | 2764 (74%) | 1427 (78%) |
| Systemic therapy | 799 (32%) | 950 (36%) | 814 (38%) | 1425 (38%) | 763 (42%) |
| Tamoxifen<sup>b</sup> | 831 (33%) | 994 (38%) | 819 (39%) | 1461 (39%) | 797 (43%) |
| Aromatase inhibitors | 1649 (66%) | 1680 (64%) | 1340 (63%) | 2381 (64%) | 1151 (63%) |

<sup>a</sup> Range given to avoid disclosure of small counts; <sup>b</sup> Medication use at anytime after breast cancer diagnosis

#### Breast cancer-specific mortality

The association between house value, deprivation and mortality are shown in Table 2 and Supplementary Figure 1. Based upon the Kaplan-Meier curves (Supplementary Figure 1), death from breast cancer at 5 years was 16.2% in patients living in the least valuable property category compared with 8.9% in patients in the most valuable category. The rate of breast cancer-specific mortality in patients in the least valuable category was 1.90 (95% CI 1.59, 2.27) times that of the most valuable category, and there was a trend across house value categories. This association was slightly attenuated after adjusting for age and comorbidities (adjusted HR=1.60 95% CI 1.34, 1.92) but further adjustment for deprivation had little impact (adjusted HR=1.63 95% CI 1.34, 1.99). The association remained after adjusting for stage and grade (adjusted HR=1.50 95% 1.20, 1.86).

**Table 2.**
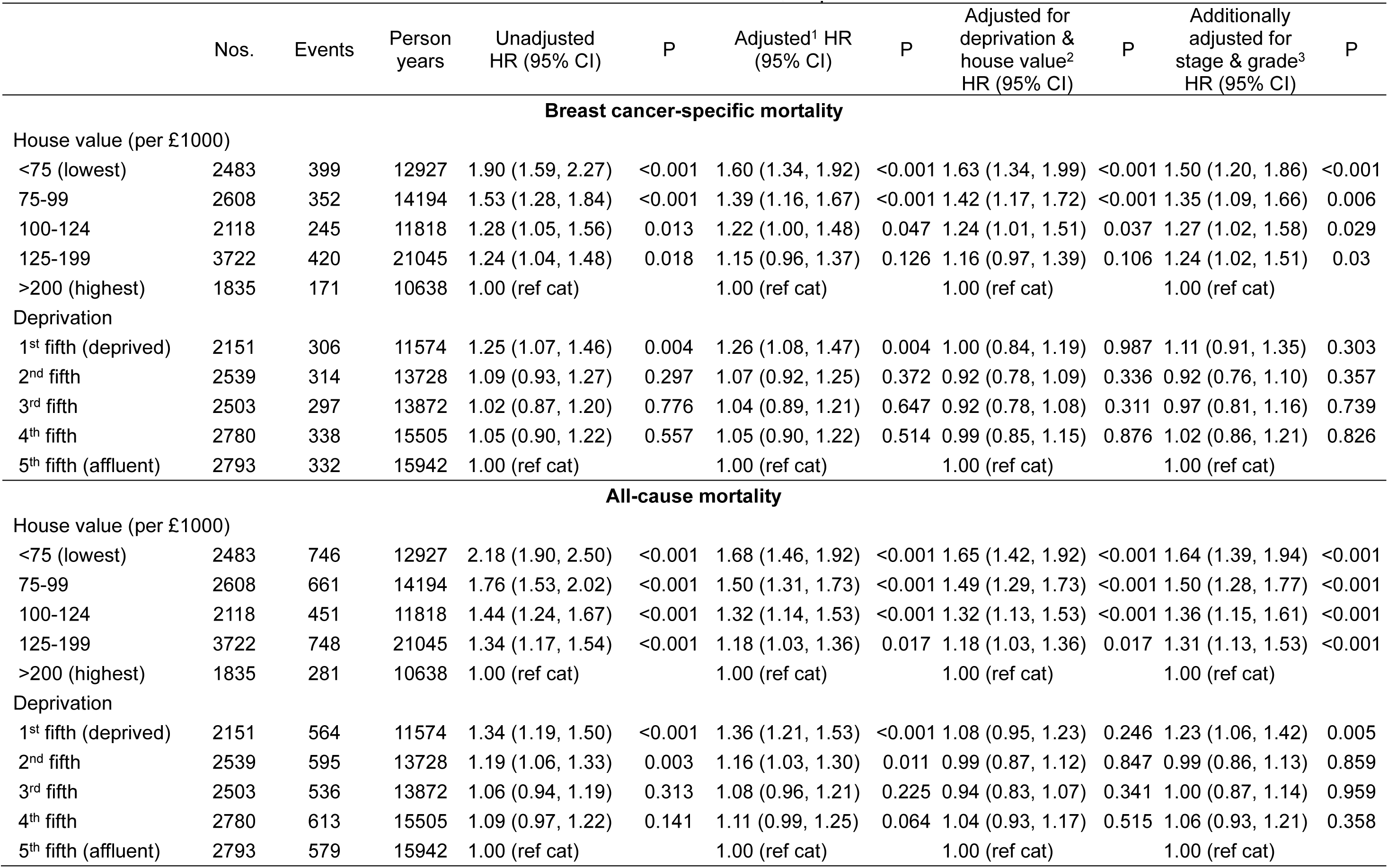
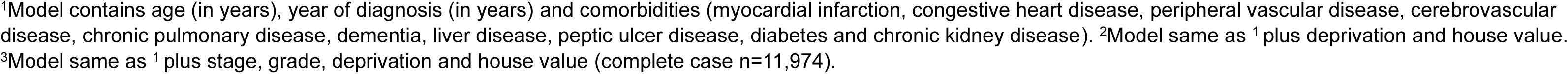
Association between house value, deprivation and survival.

|  | Nos. | Events | Person<br>years | Unadjusted<br>HR (95% CI) | P | Adjusted<sup>1</sup> HR<br>(95% CI) | P | Adjusted for<br>deprivation &<br>house value<sup>2</sup><br>HR (95% CI) | P | Additionally<br>adjusted for<br>stage & grade<sup>3</sup><br>HR (95% CI) | P |
| --- | --- | --- | --- | --- | --- | --- | --- | --- | --- | --- | --- |
| **Breast cancer-specific mortality** |  |  |  |  |  |  |  |  |  |  |  |
| House value (per £1000) |  |  |  |  |  |  |  |  |  |  |  |
| <75 (lowest) | 2483 | 399 | 12927 | 1.90 (1.59, 2.27) | <0.001 | 1.60 (1.34, 1.92) | <0.001 | 1.63 (1.34, 1.99) | <0.001 | 1.50 (1.20, 1.86) | <0.001 |
| 75-99 | 2608 | 352 | 14194 | 1.53 (1.28, 1.84) | <0.001 | 1.39 (1.16, 1.67) | <0.001 | 1.42 (1.17, 1.72) | <0.001 | 1.35 (1.09, 1.66) | 0.006 |
| 100-124 | 2118 | 245 | 11818 | 1.28 (1.05, 1.56) | 0.013 | 1.22 (1.00, 1.48) | 0.047 | 1.24 (1.01, 1.51) | 0.037 | 1.27 (1.02, 1.58) | 0.029 |
| 125-199 | 3722 | 420 | 21045 | 1.24 (1.04, 1.48) | 0.018 | 1.15 (0.96, 1.37) | 0.126 | 1.16 (0.97, 1.39) | 0.106 | 1.24 (1.02, 1.51) | 0.03 |
| >200 (highest) | 1835 | 171 | 10638 | 1.00 (ref cat) |  | 1.00 (ref cat) |  | 1.00 (ref cat) |  | 1.00 (ref cat) |  |
| Deprivation |  |  |  |  |  |  |  |  |  |  |  |
| 1<sup>st</sup> fifth (deprived) | 2151 | 306 | 11574 | 1.25 (1.07, 1.46) | 0.004 | 1.26 (1.08, 1.47) | 0.004 | 1.00 (0.84, 1.19) | 0.987 | 1.11 (0.91, 1.35) | 0.303 |
| 2<sup>nd</sup> fifth | 2539 | 314 | 13728 | 1.09 (0.93, 1.27) | 0.297 | 1.07 (0.92, 1.25) | 0.372 | 0.92 (0.78, 1.09) | 0.336 | 0.92 (0.76, 1.10) | 0.357 |
| 3<sup>rd</sup> fifth | 2503 | 297 | 13872 | 1.02 (0.87, 1.20) | 0.776 | 1.04 (0.89, 1.21) | 0.647 | 0.92 (0.78, 1.08) | 0.311 | 0.97 (0.81, 1.16) | 0.739 |
| 4<sup>th</sup> fifth | 2780 | 338 | 15505 | 1.05 (0.90, 1.22) | 0.557 | 1.05 (0.90, 1.22) | 0.514 | 0.99 (0.85, 1.15) | 0.876 | 1.02 (0.86, 1.21) | 0.826 |
| 5<sup>th</sup> fifth (affluent) | 2793 | 332 | 15942 | 1.00 (ref cat) |  | 1.00 (ref cat) |  | 1.00 (ref cat) |  | 1.00 (ref cat) |  |
| **All-cause mortality** |  |  |  |  |  |  |  |  |  |  |  |
| House value (per £1000) |  |  |  |  |  |  |  |  |  |  |  |
| <75 (lowest) | 2483 | 746 | 12927 | 2.18 (1.90, 2.50) | <0.001 | 1.68 (1.46, 1.92) | <0.001 | 1.65 (1.42, 1.92) | <0.001 | 1.64 (1.39, 1.94) | <0.001 |
| 75-99 | 2608 | 661 | 14194 | 1.76 (1.53, 2.02) | <0.001 | 1.50 (1.31, 1.73) | <0.001 | 1.49 (1.29, 1.73) | <0.001 | 1.50 (1.28, 1.77) | <0.001 |
| 100-124 | 2118 | 451 | 11818 | 1.44 (1.24, 1.67) | <0.001 | 1.32 (1.14, 1.53) | <0.001 | 1.32 (1.13, 1.53) | <0.001 | 1.36 (1.15, 1.61) | <0.001 |
| 125-199 | 3722 | 748 | 21045 | 1.34 (1.17, 1.54) | <0.001 | 1.18 (1.03, 1.36) | 0.017 | 1.18 (1.03, 1.36) | 0.017 | 1.31 (1.13, 1.53) | <0.001 |
| >200 (highest) | 1835 | 281 | 10638 | 1.00 (ref cat) |  | 1.00 (ref cat) |  | 1.00 (ref cat) |  | 1.00 (ref cat) |  |
| Deprivation |  |  |  |  |  |  |  |  |  |  |  |
| 1<sup>st</sup> fifth (deprived) | 2151 | 564 | 11574 | 1.34 (1.19, 1.50) | <0.001 | 1.36 (1.21, 1.53) | <0.001 | 1.08 (0.95, 1.23) | 0.246 | 1.23 (1.06, 1.42) | 0.005 |
| 2<sup>nd</sup> fifth | 2539 | 595 | 13728 | 1.19 (1.06, 1.33) | 0.003 | 1.16 (1.03, 1.30) | 0.011 | 0.99 (0.87, 1.12) | 0.847 | 0.99 (0.86, 1.13) | 0.859 |
| 3<sup>rd</sup> fifth | 2503 | 536 | 13872 | 1.06 (0.94, 1.19) | 0.313 | 1.08 (0.96, 1.21) | 0.225 | 0.94 (0.83, 1.07) | 0.341 | 1.00 (0.87, 1.14) | 0.959 |
| 4<sup>th</sup> fifth | 2780 | 613 | 15505 | 1.09 (0.97, 1.22) | 0.141 | 1.11 (0.99, 1.25) | 0.064 | 1.04 (0.93, 1.17) | 0.515 | 1.06 (0.93, 1.21) | 0.358 |
| 5<sup>th</sup> fifth (affluent) | 2793 | 579 | 15942 | 1.00 (ref cat) |  | 1.00 (ref cat) |  | 1.00 (ref cat) |  | 1.00 (ref cat) |  |

<sup>1</sup>Model contains age (in years), year of diagnosis (in years) and comorbidities (myocardial infarction, congestive heart disease, peripheral vascular disease, cerebrovascular disease, chronic pulmonary disease, dementia, liver disease, peptic ulcer disease, diabetes and chronic kidney disease). <sup>2</sup>Model same as <sup>1</sup> plus deprivation and house value. <sup>3</sup>Model same as <sup>1</sup> plus stage, grade, deprivation and house value (complete case n=11,974).

Based upon the Kaplan-Meier curves, death from breast cancer at 5 years was 14.8% in patients living in the most deprived areas compared with 12.1% in the least deprived areas. The rate of breast cancer-specific mortality in patients in the most deprived areas was 1.25 (95% CI 1.07, 1.46) times that of the least deprived areas. This association was similar after adjusting for age and comorbidities (adjusted HR=1.26 95% CI 1.08, 1.47) but was attenuated after further adjustment for house value (adjusted HR=1.00 95% CI 0.84, 1.19) and stage and grade (adjusted HR=1.11 95% 0.91, 1.35). Similar patterns of association were observed for all-cause mortality.

#### Stage of disease at diagnosis

The association between house value, deprivation and late stage at diagnosis is shown in Table 3. Overall, in breast cancer patients there was evidence of later-stage diagnosis in those living in the least valuable properties, with 7.5% of patients in the least valuable house category diagnosed with stage 4 breast cancer compared with 4.1% of patients in the most valuable category (OR=1.89 95% CI 1.43, 2.50). This association persisted after adjusting for age, year of diagnosis and comorbidities (adjusted OR=1.65 95% CI 1.24, 2.20) and for deprivation (adjusted OR=1.70 95% CI 1.24, 2.32). Similarly, 5.9% of patients from the most deprived areas had stage 4 breast cancer compared with 5.0% of patients from the least deprived areas (OR=1.20 95% CI 0.93, 1.54). This association was similar after adjustments for age, year of diagnosis and comorbidities (adjusted OR=1.21 95% CI 0.94, 1.57) but was attenuated after adjusting for house value (adjusted OR=0.92 95% CI 0.69, 1.23). The associations were slightly less marked when late stage was expanded to include both stage 4 or stage unknown (after adjusting for age, year and comorbidities; house value adjusted OR=1.34 95% CI 1.07-1.67); deprivation OR=1.20 95% CI 0.98-1.47).

**Table 3.**
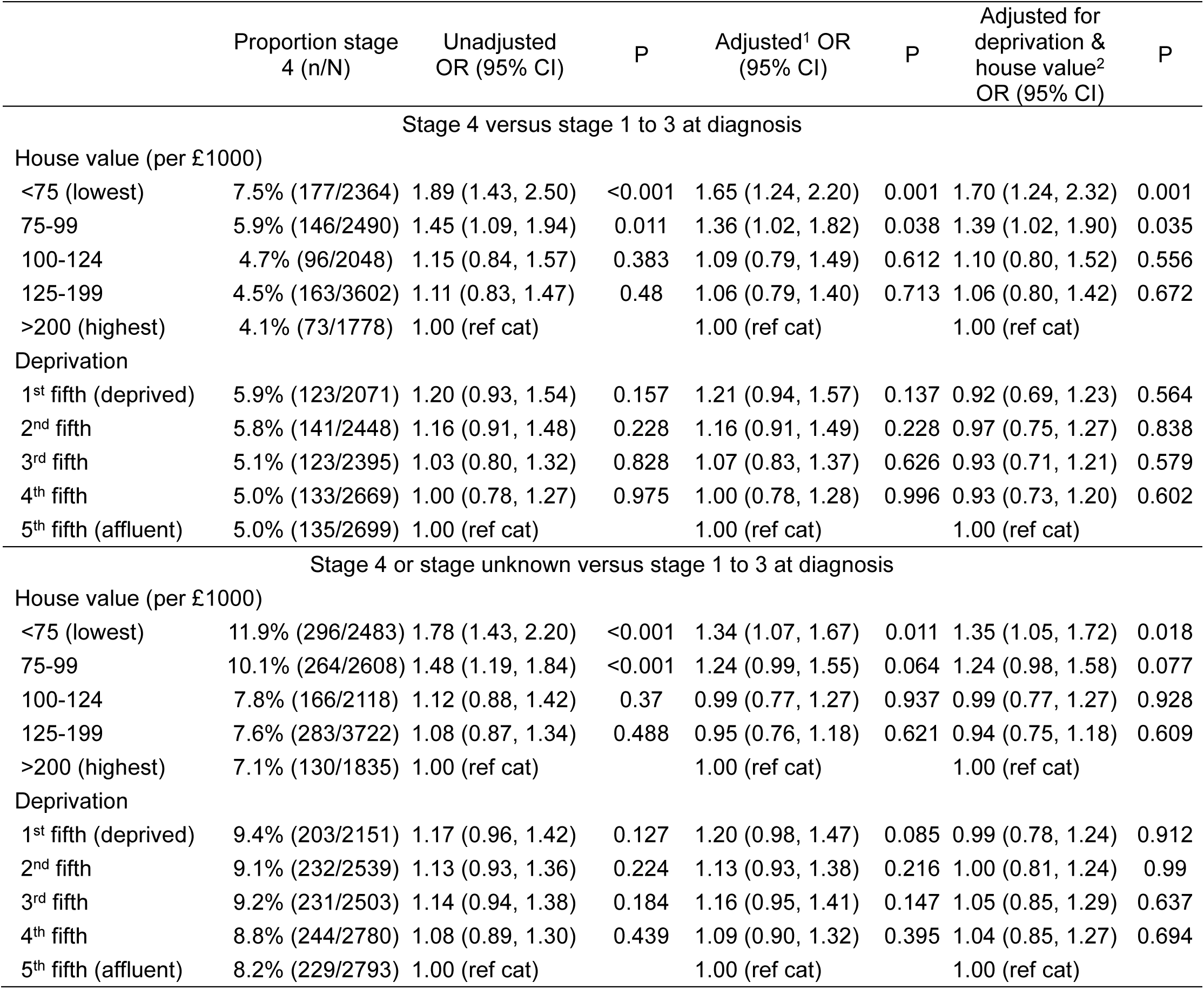
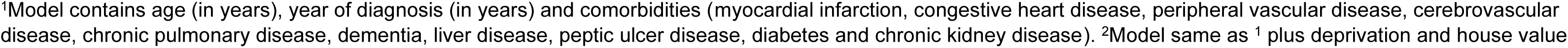
Association between house value and proportion of stage 4 breast cancer diagnoses.

|  |  |  |  |  |  |  |  |
| --- | --- | --- | --- | --- | --- | --- | --- |
|  | Proportion stage<br>4 (n/N) | Unadjusted<br>OR (95% CI) | P | Adjusted<sup>1</sup> OR<br>(95% CI) | P | Adjusted for<br>deprivation &<br>house value<sup>2</sup><br>OR (95% CI) | P |
| Stage 4 versus stage 1 to 3 at diagnosis |  |  |  |  |  |  |  |
| House value (per £1000) |  |  |  |  |  |  |  |
| <75 (lowest) | 7.5% (177/2364) | 1.89 (1.43, 2.50) | <0.001 | 1.65 (1.24, 2.20) | 0.001 | 1.70 (1.24, 2.32) | 0.001 |
| 75-99 | 5.9% (146/2490) | 1.45 (1.09, 1.94) | 0.011 | 1.36 (1.02, 1.82) | 0.038 | 1.39 (1.02, 1.90) | 0.035 |
| 100-124 | 4.7% (96/2048) | 1.15 (0.84, 1.57) | 0.383 | 1.09 (0.79, 1.49) | 0.612 | 1.10 (0.80, 1.52) | 0.556 |
| 125-199 | 4.5% (163/3602) | 1.11 (0.83, 1.47) | 0.48 | 1.06 (0.79, 1.40) | 0.713 | 1.06 (0.80, 1.42) | 0.672 |
| >200 (highest) | 4.1% (73/1778) | 1.00 (ref cat) |  | 1.00 (ref cat) |  | 1.00 (ref cat) |  |
| Deprivation |  |  |  |  |  |  |  |
| 1<sup>st</sup> fifth (deprived) | 5.9% (123/2071) | 1.20 (0.93, 1.54) | 0.157 | 1.21 (0.94, 1.57) | 0.137 | 0.92 (0.69, 1.23) | 0.564 |
| 2<sup>nd</sup> fifth | 5.8% (141/2448) | 1.16 (0.91, 1.48) | 0.228 | 1.16 (0.91, 1.49) | 0.228 | 0.97 (0.75, 1.27) | 0.838 |
| 3<sup>rd</sup> fifth | 5.1% (123/2395) | 1.03 (0.80, 1.32) | 0.828 | 1.07 (0.83, 1.37) | 0.626 | 0.93 (0.71, 1.21) | 0.579 |
| 4<sup>th</sup> fifth | 5.0% (133/2669) | 1.00 (0.78, 1.27) | 0.975 | 1.00 (0.78, 1.28) | 0.996 | 0.93 (0.73, 1.20) | 0.602 |
| 5<sup>th</sup> fifth (affluent) | 5.0% (135/2699) | 1.00 (ref cat) |  | 1.00 (ref cat) |  | 1.00 (ref cat) |  |
| Stage 4 or stage unknown versus stage 1 to 3 at diagnosis |  |  |  |  |  |  |  |
| House value (per £1000) |  |  |  |  |  |  |  |
| <75 (lowest) | 11.9% (296/2483) | 1.78 (1.43, 2.20) | <0.001 | 1.34 (1.07, 1.67) | 0.011 | 1.35 (1.05, 1.72) | 0.018 |
| 75-99 | 10.1% (264/2608) | 1.48 (1.19, 1.84) | <0.001 | 1.24 (0.99, 1.55) | 0.064 | 1.24 (0.98, 1.58) | 0.077 |
| 100-124 | 7.8% (166/2118) | 1.12 (0.88, 1.42) | 0.37 | 0.99 (0.77, 1.27) | 0.937 | 0.99 (0.77, 1.27) | 0.928 |
| 125-199 | 7.6% (283/3722) | 1.08 (0.87, 1.34) | 0.488 | 0.95 (0.76, 1.18) | 0.621 | 0.94 (0.75, 1.18) | 0.609 |
| >200 (highest) | 7.1% (130/1835) | 1.00 (ref cat) |  | 1.00 (ref cat) |  | 1.00 (ref cat) |  |
| Deprivation |  |  |  |  |  |  |  |
| 1<sup>st</sup> fifth (deprived) | 9.4% (203/2151) | 1.17 (0.96, 1.42) | 0.127 | 1.20 (0.98, 1.47) | 0.085 | 0.99 (0.78, 1.24) | 0.912 |
| 2<sup>nd</sup> fifth | 9.1% (232/2539) | 1.13 (0.93, 1.36) | 0.224 | 1.13 (0.93, 1.38) | 0.216 | 1.00 (0.81, 1.24) | 0.99 |
| 3<sup>rd</sup> fifth | 9.2% (231/2503) | 1.14 (0.94, 1.38) | 0.184 | 1.16 (0.95, 1.41) | 0.147 | 1.05 (0.85, 1.29) | 0.637 |
| 4<sup>th</sup> fifth | 8.8% (244/2780) | 1.08 (0.89, 1.30) | 0.439 | 1.09 (0.90, 1.32) | 0.395 | 1.04 (0.85, 1.27) | 0.694 |
| 5<sup>th</sup> fifth (affluent) | 8.2% (229/2793) | 1.00 (ref cat) |  | 1.00 (ref cat) |  | 1.00 (ref cat) |  |

<sup>1</sup>Model contains age (in years), year of diagnosis (in years) and comorbidities (myocardial infarction, congestive heart disease, peripheral vascular disease, cerebrovascular disease, chronic pulmonary disease, dementia, liver disease, peptic ulcer disease, diabetes and chronic kidney disease). <sup>2</sup>Model same as <sup>1</sup> plus deprivation and house value

Table 4 contains sensitivity analyses that address individuals with missing house value or deprivation, using a separate missing indicator category or multiple imputation. These analyses were generally similar to the main findings; however, individuals with missing house valuations had substantially higher breast cancer-specific mortality (HR=2.93 95% CI 2.40, 3.57).

**Table 4.** Sensitivity analyses for the association between house value and breast cancer-specific survival.

| Outcome | Nos. | Breast cancer deaths | Person Years | Unadjusted HR (95% CI) | P | Adjusted<sup>1</sup> HR (95% CI) | P | Adjusted for deprivation & house value<sup>2</sup> HR (95% CI) | P |
| --- | --- | --- | --- | --- | --- | --- | --- | --- | --- |
| A. Including missing house value and deprivation as separate category |  |  |  |  |  |  |  |  |  |
| House value (per £1000) |  |  |  |  |  |  |  |  |  |
| <75 (lowest) | 2504 | 403 | 13036 | 1.88 (1.57, 2.25) | <0.001 | 1.56 (1.31, 1.87) | <0.001 | 1.58 (1.30, 1.92) | <0.001 |
| 75-99 | 2630 | 354 | 14329 | 1.51 (1.26, 1.81) | <0.001 | 1.36 (1.14, 1.64) | 0.001 | 1.39 (1.15, 1.68) | 0.001 |
| 100-124 | 2136 | 247 | 11899 | 1.27 (1.05, 1.54) | 0.015 | 1.20 (0.99, 1.46) | 0.063 | 1.22 (1.00, 1.49) | 0.049 |
| 125-199 | 3748 | 421 | 21207 | 1.22 (1.02, 1.46) | 0.027 | 1.13 (0.94, 1.35) | 0.185 | 1.14 (0.95, 1.36) | 0.155 |
| >200 (highest) | 1852 | 174 | 10738 | 1.00 (ref cat) |  | 1.00 (ref cat) |  | 1.00 (ref cat) |  |
| Missing | 976 | 221 | 4608 | 2.93 (2.40, 3.57) | <0.001 | 2.25 (1.84, 2.76) | <0.001 | 2.31 (1.88, 2.85) | <0.001 |
| Deprivation |  |  |  |  |  |  |  |  |  |
| 1<sup>st</sup> fifth (deprived) | 2298 | 342 | 12285 | 1.22 (1.05, 1.41) | 0.008 | 1.24 (1.07, 1.43) | 0.005 | 1.00 (0.85, 1.18) | 0.978 |
| 2<sup>nd</sup> fifth | 2741 | 360 | 14733 | 1.07 (0.93, 1.24) | 0.35 | 1.07 (0.93, 1.24) | 0.359 | 0.92 (0.79, 1.07) | 0.296 |
| 3<sup>rd</sup> fifth | 2705 | 343 | 14931 | 1.02 (0.88, 1.18) | 0.823 | 1.04 (0.90, 1.20) | 0.624 | 0.91 (0.78, 1.06) | 0.243 |
| 4<sup>th</sup> fifth | 2979 | 377 | 16428 | 1.02 (0.88, 1.17) | 0.819 | 1.03 (0.89, 1.19) | 0.679 | 0.96 (0.83, 1.11) | 0.562 |
| 5<sup>th</sup> fifth (affluent) | 2938 | 371 | 16433 | 1.00 (ref cat) |  | 1.00 (ref cat) |  | 1.00 (ref cat) |  |
| Missing | 185 | 27 | 1006 | 1.20 (0.81, 1.78) | 0.351 | 1.23 (0.83, 1.82) | 0.303 | 0.84 (0.56, 1.25) | 0.393 |
| B. Using multiple imputation to impute house value and deprivation<sup>3</sup> |  |  |  |  |  |  |  |  |  |
| House value (per £1000) |  |  |  |  |  |  |  |  |  |
| <75 (lowest) | 2731 | 469 | 14048 | 1.90 (1.60, 2.25) | <0.001 | 1.58 (1.33, 1.87) | <0.001 | 1.61 (1.33, 1.95) | <0.001 |
| 75-99 | 2850 | 404 | 15353 | 1.54 (1.30, 1.83) | <0.001 | 1.39 (1.17, 1.65) | <0.001 | 1.42 (1.18, 1.70) | <0.001 |
| 100-124 | 2276 | 282 | 12588 | 1.29 (1.06, 1.57) | 0.01 | 1.22 (1.00, 1.49) | 0.049 | 1.24 (1.01, 1.52) | 0.036 |
| 125-199 | 4019 | 471 | 22529 | 1.21 (1.02, 1.45) | 0.031 | 1.12 (0.94, 1.34) | 0.202 | 1.13 (0.95, 1.35) | 0.167 |
| >200 (highest) | 1970 | 194 | 11299 | 1.00 (ref cat) |  | 1.00 (ref cat) |  | 1.00 (ref cat) |  |
| Deprivation |  |  |  |  |  |  |  |  |  |
| 1<sup>st</sup> fifth (deprived) | 2332 | 347 | 12455 | 1.22 (1.05, 1.41) | 0.008 | 1.23 (1.06, 1.43) | 0.005 | 0.98 (0.83, 1.16) | 0.832 |
| 2<sup>nd</sup> fifth | 2772 | 368 | 14882 | 1.07 (0.93, 1.24) | 0.341 | 1.07 (0.93, 1.24) | 0.355 | 0.92 (0.79, 1.07) | 0.286 |
| 3<sup>rd</sup> fifth | 2741 | 347 | 15133 | 1.01 (0.88, 1.17) | 0.85 | 1.03 (0.89, 1.20) | 0.652 | 0.92 (0.79, 1.07) | 0.267 |
| 4<sup>th</sup> fifth | 3024 | 380 | 16671 | 1.01 (0.88, 1.17) | 0.873 | 1.02 (0.89, 1.18) | 0.74 | 0.96 (0.83, 1.11) | 0.598 |
| 5<sup>th</sup> fifth (affluent) | 2977 | 378 | 16677 | 1.00 (ref cat) |  | 1.00 (ref cat) |  | 1.00 (ref cat) |  |

<sup>1</sup>Model contains age (in years), year of diagnosis (in years) and comorbidities (myocardial infarction, congestive heart disease, peripheral vascular disease, cerebrovascular disease, chronic pulmonary disease, dementia, liver disease, peptic ulcer disease, diabetes and chronic kidney disease). <sup>2</sup>Model same as <sup>1</sup> plus deprivation and house value. <sup>3</sup>Number of women, breast cancer deaths and person years based upon first imputed dataset.

## Discussion

In this population-based study of 12,766 women diagnosed with breast cancer in NI, we observed associations between both house value and deprivation and breast-cancer specific mortality and stage 4 disease presentation. However, the observed associations were much more pronounced for house value, an individual-level measure of SES, than the more widely used area-based NIMDM. For example, women living in the lowest valued properties had a 60% increase in breast cancer-specific mortality compared with women living in the highest valued properties, and this association persisted after further adjustment for deprivation. In contrast, women in the most deprived areas had a 26% increase in breast cancer-specific mortality compared with women in the least deprived areas, with the association substantially attenuated after adjustment for house value, suggesting that individual-level SES measures may capture socio-economic influences on breast cancer outcomes more precisely than area-based measures.

Our study’s findings of increased breast cancer-specific mortality among women with lower SES are consistent with previous research demonstrating socio-economic gradients in breast cancer outcomes [5, 30–32]. Few studies have specifically examined house value in relation to breast cancer survival, but of those that did, evidence supports our findings. A Singaporean study observed a 63% increase in cancer-specific mortality in women living in the lowest versus highest property value categories, though this was attenuated to 14% after adjusting for confounders including tumour stage [19]. Similarly, a USA study demonstrated a reduction in breast cancer-specific mortality with increasing house value [20]. Our study observed a 60% increase in mortality for the lowest versus highest house value categories, which remained substantial even after adjustment for tumour characteristics.

Previous studies using other individual measures of SES, such as income, occupation, and education, have generally demonstrated poorer breast cancer survival among lower SES groups. A systematic review of studies investigating individual socio-economic measures found occupation, income, and education were associated with poorer breast cancer survival [6], though none of the included studies were UK-based. A previous UK study investigating census-based individual-level socio-economic measures showed limited evidence of associations between education or occupation and 5-year net survival, but found improved survival among individuals with the highest income estimated from occupation [5].

Direct comparison with these studies is challenging as we used house value as our individual-level SES measure. However, house value may provide clearer indications of socio-economic inequalities than other measures. The strong socio-economic gradient we observed could reflect the high levels of deprivation in NI [25, 33] and/or the long waiting lists for health care, which may exacerbate inequalities if more affluent women can access private healthcare to overcome delays. Data on private healthcare utilisation were not available precluding further investigation of this mechanism.

Our findings also suggest that the individual-level SES indicators may be more sensitive than area-level measures for detecting socio-economic differences in breast cancer-specific mortality in NI. Area-based analyses may dilute true SES effects because all individuals within a specific area are assigned the same average deprivation level regardless of their household circumstances [12]. This limitation may be particularly relevant for the NIMDM which uses Super Output Areas which averages approximately 2,100 people, potentially masking individual-level socio-economic variation.

The mechanisms underlying the observed socio-economic inequalities are likely multifactorial. One explanation may be the differences in rates of breast cancer screening, with those more socially disadvantaged less likely to attend breast screening and therefore more likely to be diagnosed with advanced disease and have poorer outcomes [34, 35]. Moreover, prior research has demonstrated that Black women have poorer breast cancer survival than White women, which is partly attributable to more aggressive tumour characteristics [36, 37]. While NI has substantially less ethnic diversity than other UK regions, it exhibits higher levels of social deprivation [33, 38]. Previous studies suggest that patients from lower socio-economic backgrounds may have different tumour characteristics, including later stage disease [39, 40]. Indeed, we observed evidence of later-stage disease presentation among patients with lower house values, although associations with mortality persisted after adjustments for stage, suggesting additional pathways beyond diagnostic timing. Furthermore, patients from lower SES backgrounds may experience healthcare differently, including altered referral patterns and treatment provision [39].

Patients from lower socio-economic backgrounds may experience poorer health and more behavioural risk factors, including weight, diet, alcohol intake and exercise, which could impact cancer outcomes. However, we did not capture any data on lifestyle characteristics. Furthermore, lower health literacy, particularly prevalent among deprived populations, may limit breast health awareness and symptom recognition, potentially contributing to diagnostic delays and reduced screening participation [4].Further research investigating healthcare, lifestyle factors, and other individual-level SES measures in NI (such as education, occupation, transport access, and disability status) is needed to better understand the mechanisms underlying the association between house value and breast cancer outcomes

### Strengths and limitations

The main strength of our study is the use of a large, well-characterised population-based cohort of breast cancer patients with comprehensive coverage of domestic property valuations [14].

Our study has various limitations. House value data was missing for 7% of breast cancer patients because the address did not match a UPRN. This may reflect incorrect address or properties without a domestic valuation (e.g., a residential care setting, those with no fixed address), which could explain the higher mortality observed in patients with missing house values. The house values in our analysis were used to allocate local tax in 2025, but they are based upon what the house could have reasonably sold for in 2005 and over the last two decades absolute house prices have considerably changed. However, this approach should not introduce bias provided the relative socio-economic ranking of property values across different areas remained stable over time, even when absolute property values change, as the measure captures relative rather than absolute wealth position within a population. House value may not adequately reflect the resident’s current financial position as their wealth may be tied up in assets, but they have limited disposable income. Additionally, we lacked data on home ownership. While over 70% of homes among persons aged 65 years and older in NI are owner-occupied[14], house value may still serve as a reasonable SES marker for private renters, and public sector tenants are likely to rent properties in lower value categories. Nevertheless, there is potential for exposure misclassification between two individuals occupying houses of a similar value, if one is a tenant, and the other a homeowner, due to the large wealth gap between them. We could not adjust for clustering as we only had access to the NIMDM quintile and not the Super Output Area of the patient’s address. This may have led to an overestimation of statistical power and potentially inflated the precision of our estimates related to deprivation. However, this should not impact upon the individual-level analysis of house value. Finally, we only investigated breast cancer, which has a widely publicised and used screening programme. Further research is needed to determine whether house value serves as a useful indicator of health inequalities for other cancer sites.

In summary, both house value and area-level deprivation measures were associated with stage 4 disease presentation and breast cancer-specific mortality. However, associations were more pronounced for house value, suggesting that individual-level SES measures may serve as more sensitive indicators for monitoring socio-economic inequalities in breast cancer outcomes at the population level. Further research is warranted to better understand the mechanisms underlying these inequalities and to inform targeted interventions aimed at reducing socio-economic disparities in breast cancer outcomes.

## Data Availability

The dataset can be recreated by authorised researchers, at a cost, after obtaining the necessary ethical approvals and applying to the Northern Ireland Cancer Registry. Further information on how to apply can be found here: https://www.qub.ac.uk/research-centres/nicr/CancerInformation/requests/. Analysis codes are available upon request to the corresponding author.

## Acknowledgements

This work would not have been possible without the expertise and dedication of NI Cancer Registry personnel including Cancer Intelligence Officers, IT, administrative and management staff. This work uses data provided by patients and collected by Health and Social Care NI as part of their care and support. The NI Cancer Registry is funded by the Public Health Agency of NI. The authors alone are responsible for the interpretation of the data. Any views or opinions presented are solely those of the authors.

## Author Contributions

**Sarah M. Baxter:** Writing -review & editing**; Charlene M. Mc Shane:** Conceptualisation, Funding acquisition, Methodology, Writing -review & editing**; Stuart A. McIntosh:** Conceptualisation, Funding acquisition, Methodology, Writing -review & editing**; Damien Bennett:** Conceptualisation, Funding acquisition, Data Duration, Methodology, Writing – review & editing; **Meenakshi Sharma:** Writing – review & editing; **Lynne Lohfeld**: Conceptualisation, Funding acquisition, Methodology, Writing -review & editing**; Daniel R.S. Middleton**: Funding acquisition, Writing -review & editing: **Gerard Savage**: Data curation, Writing – review & editing; **Deirdre Fitzpatrick**: Data curation, Writing – review & editing; **Ann Mc Brien:** Conceptualisation, Funding acquisition, Methodology, Writing -review & editing; **David McCallion**: Writing -review & editing**; Anna Gavin**: Conceptualisation, Funding acquisition, Methodology, Writing -review & editing**; Chris R. Cardwell**: Conceptualisation, Funding acquisition, Software, Validation, Formal analysis, Visualisation; Supervision, Project administration, Methodology, Writing – Original draft, Writing -review & editing.

## Declaration of generative AI and AI-assisted technologies in the writing process

During the preparation of this work the authors used Claude AI to improve grammar and sentence structure. After using this tool/service, the authors reviewed and edited the content as needed and take full responsibility for the content of the published article.

