## Supplementary Figure 1 for "House value as an individual socioeconomic indicator for breast cancer survival and late-stage diagnosis: a population-based cohort study from Northern Ireland"

Supplementary Figure 1. Kaplan-Meier survival curves for breast cancer-specific and all-cause mortality by house value and deprivation.

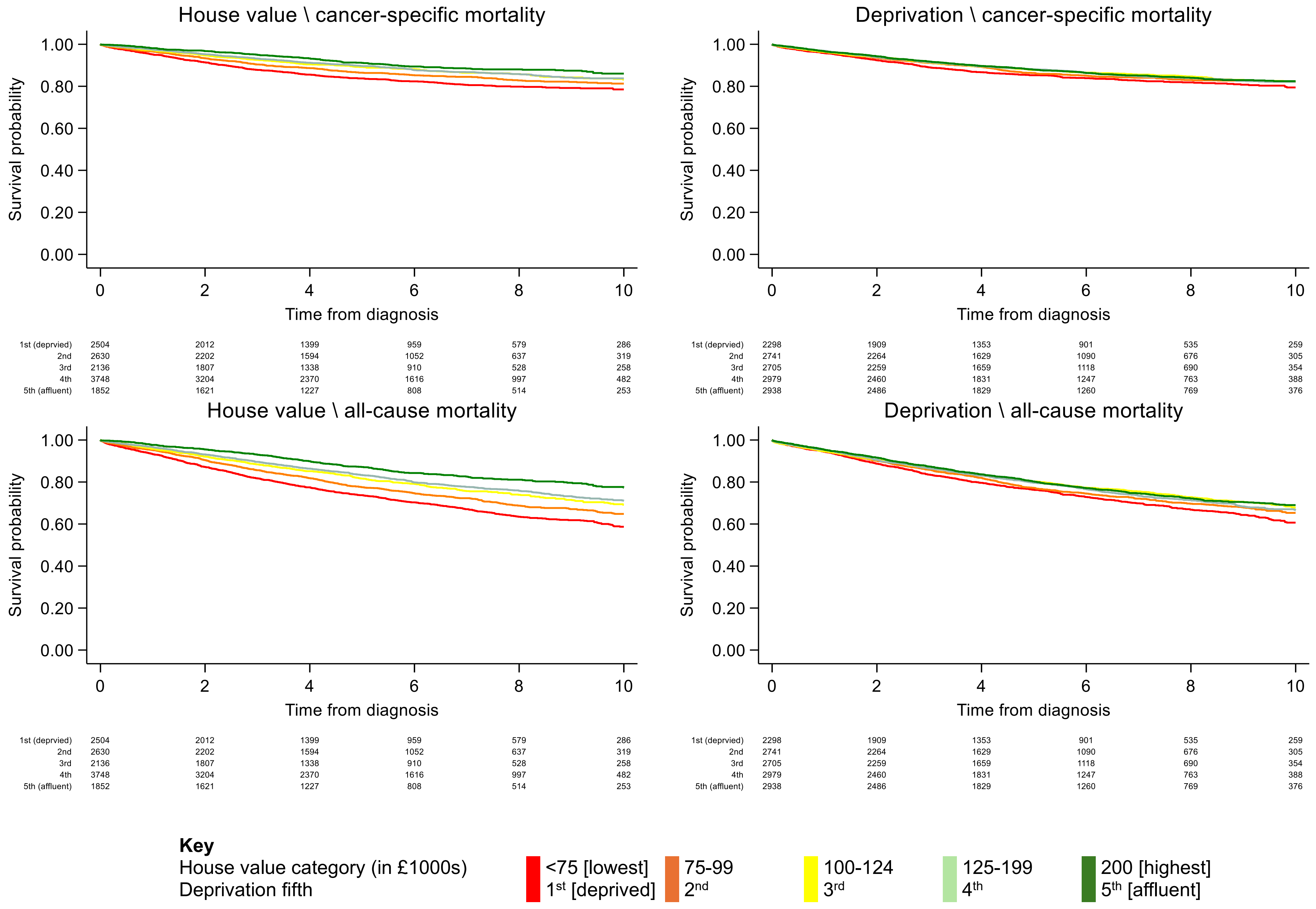
